# Outcomes of COVID-19 among Patients with End Stage Renal Disease on Remdesivir

**DOI:** 10.1101/2021.02.10.21251527

**Authors:** Vijairam Selvaraj, Muhammad Baig, Kwame Dapaah-Afriyie, Arkadiy Finn, Atin Jindal, George Bayliss

**Affiliations:** Division of Hospital Medicine, The Miriam Hospital, Providence, Rhode Island; Warren Alpert Medical School of Brown University, Providence, Rhode Island; Division of Kidney Disease and Hypertension, Rhode Island Hospital, Providence, Rhode Island

**Keywords:** COVID-19, remdesivir, end stage renal disease, hemodialysis

## Abstract

**BACKGROUND:** Since the beginning of the COVID-19 pandemic, there has been widespread use of remdesivir in adults and children. There is little known information about its outcomes in patients with severe renal dysfunction or end-stage renal disease who are on hemodialysis.

**METHODS:** A retrospective, multicenter study was conducted on patients with end-stage renal disease on hemodialysis that were discharged after treatment for COVID-19 between April 1st and December 31st, 2020. Primary endpoints were the length of stay, mortality, maximum oxygen requirements along with the escalation of care needing mechanical ventilation. Secondary endpoints included change in C reactive protein, d dimer levels, and disposition.

**RESULTS:** A total of 52 charts were reviewed, of which 28 met the inclusion criteria. 14 patients received remdesivir, and 14 patients did not receive remdesivir. The majority of patients were caucasian, female, with diabetes mellitus and hypertension. The mean age was 65.33 +14.14 years. All the patients in the remdesivir group received dexamethasone as compared to only 30% of patients in the non-remdesivir group. There was no significant difference in C reactive protein, d dimer levels, and disposition between the two groups. Approximately 35% of the patients died, 18% required intensive ventilation, and the mean length of stay was 12.21 days.

**DISCUSSION:** The study demonstrated no clinically significant difference in length of stay, maximum oxygen requirements, or mortality in COVID-19 patients with end-stage renal disease in the remdesivir group as compared to the non-remdesivir group. Further studies are needed to study the effects of remdesivir on the renal function and disease course in patients with chronic kidney disease stage 4 or 5 that are not on dialysis.

## INTRODUCTION

COVID-19, a clinical syndrome caused by infection with the SARS-CoV2 virus, a novel coronavirus, has led to hospitalization in many cases. Remdesivir, an adenosine analog antiviral drug that inhibits viral RNA polymerase, has demonstrated in vitro activity against viruses such as Ebola, Middle East Respiratory Syndrome-Coronavirus (MERS-CoV), and Severe Acute Respiratory Syndrome-Coronavirus (SARS-CoV).

The Adaptive COVID-19 Treatment Trial-1 (ACTT-1) showed that the median time to recovery was 10 days in the Remdesivir group compared to 15 days in the placebo group. There was uncertainty as to whether Remdesivir provides a survival benefit in addition to accelerated time to recovery [1]. The Infectious Diseases Society of America panel recommended remdesivir to treat severe COVID-19 in hospitalized patients with SpO2 ≤94% on room air, including patients on supplemental oxygen or mechanical ventilation [2]. The World Health Organization issued a ‘weak or conditional recommendation against’ using remdesivir in hospitalized COVID-19 patients [3]. There is widespread use of remdesivir in COVID-19 patients. Most of the clinical trials excluded patients with CrCl<30 ml/min/1.73m^2^. There is little known information about outcomes in patients with severe renal dysfunction or end-stage renal disease (ESRD) who are on hemodialysis (HD).

Because Remdesivir has limited water solubility, the intravenous preparation contains the vehicle sulfobutylether-β-cyclodextrin (SBECD). Continuous renal replacement therapy and hemodialysis readily remove SBECD, and significant accumulation only occurs in patients when dialysis is held for prolonged periods. Concerns about the SBECD carrier’s accumulation should be allayed by the available safety data in patients with kidney failure treated with voriconazole, which uses the same carrier [4].

We postulate that the addition of Remdesivir to Dexamethasone as part of the treatment regimen in patients with COVID-19 and ESRD may reduce the length of stay and improve clinical outcomes. We report the clinical characteristics, early outcomes, and C-reactive protein (CRP) and d dimer levels in a limited set of patients with COVID-19 and ESRD on HD.

## MATERIALS AND METHODS

This was a retrospective, multicenter cohort study of patients with ESRD on HD that were discharged after treatment for COVID-19 between April 1st and December 31st, 2020. All patients tested positive for SARS-CoV2 by quantitative RT-PCR in nasopharyngeal swabs. The Institutional Review Board of the Hospital approved the study.

Dexamethasone 6mg once daily was administered to patients with COVID-19 for 10 days from July 2020 onwards based on the RECOVERY trial [5]. In patients that received remdesivir, they were given 200mg on day one, followed by 100mg once daily for four more days. Remdesivir was discontinued if there was transaminitis, with levels greater than ten times the upper limit of normal. Prone positioning was instituted as per hospital protocol in all patients with oxygen requirement > 4L/min if they could tolerate it. Patients also received antibiotics if there was a concern for superimposed bacterial infection. Select patients were also enrolled in monoclonal antibody trial if they were found eligible by the Infectious Diseases Department.

Physicians in the Division of Hospital Medicine at Miriam Hospital (an affiliate of Warren Alpert Medical School of Brown University) collected data on all discharged patients above the age of 18 with a diagnosis of COVID-19 and ESRD on HD between April 1st and December 31st, 2020. The following patients were excluded:

1. Patients < 18 years of age
2. Patients who tested positive for SARS-CoV2 but had no clinical evidence of COVID-19 or elevation in inflammatory markers.
3. Patients with ESRD who received renal transplant and are not on dialysis.
4. Patients with transaminitis, with levels greater than ten times the upper limit of normal.

**Figure 1:**
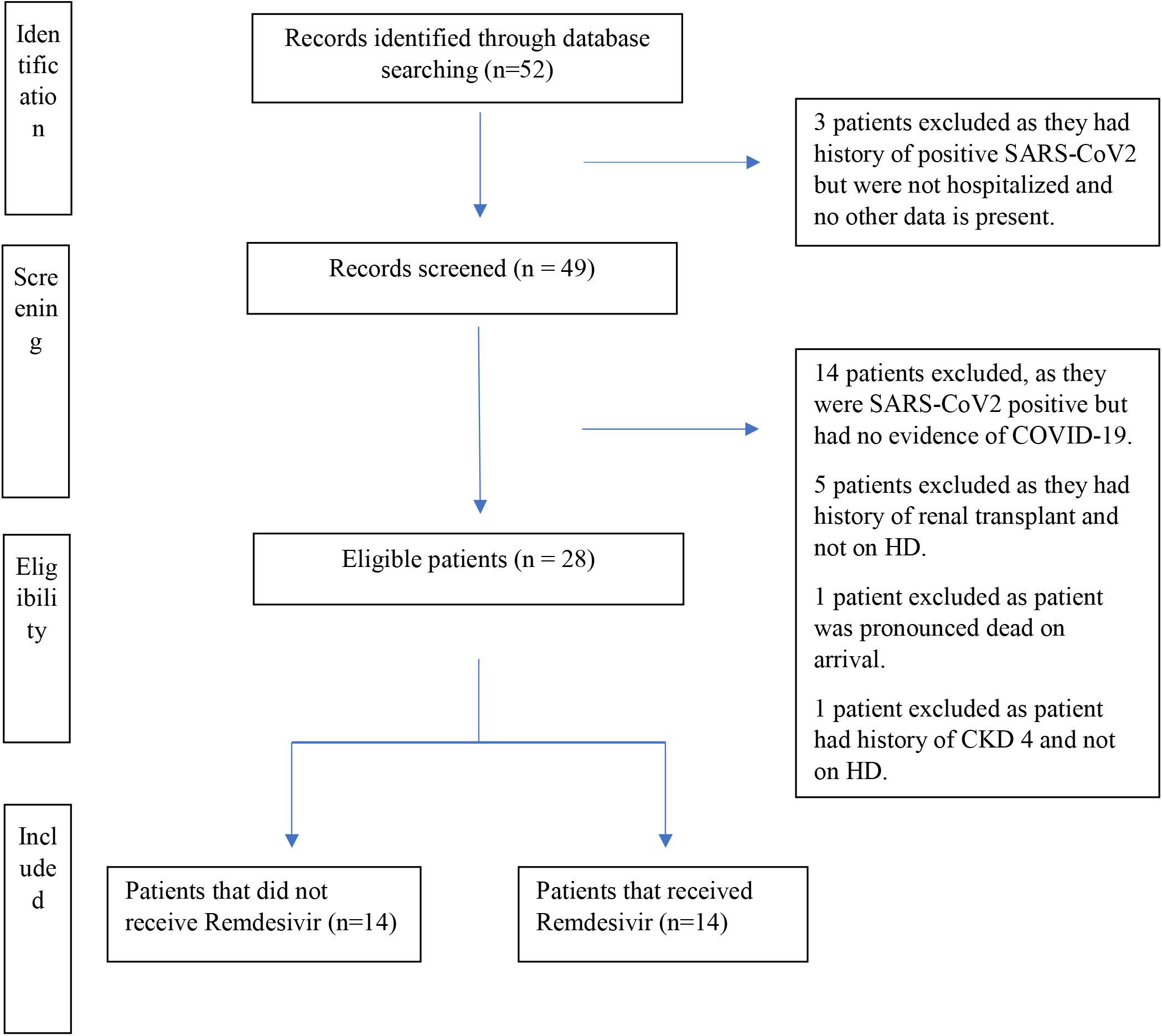
Flow Chart outlining chart review.

The Nephrology service at Miriam Hospital (an affiliate of Alpert Medical School of Brown University) followed these patients while they were admitted.

### Endpoints

Our primary endpoint was the length of stay, mortality, and maximum oxygen requirements, along with the escalation of care needing mechanical ventilation. Secondary endpoints included change in CRP levels, d dimer levels, and disposition.

### Data Collection

Data were obtained from the Epic Electronic Medical Record system and recorded in a standardized form. Demographic data, laboratory findings, maximum oxygen requirements in Liters Per Minute (LPM), length of stay (LOS), CRP levels, d dimer levels, and comorbid conditions were ascertained.

### Analysis

A two-sample t-test was used to compare means. Z-test and chi-square analysis were used to compare proportions. Type 1 error (alpha) was set at 0.05. Statistical analysis was performed using ‘R’ programming software. Demographic risk factors modeled included age, sex, race, cardiovascular disease/peripheral vascular disease, history of arrhythmia, anticoagulation, chronic lung disease, diabetes, hypertension, obesity, immunosuppression, and tobacco use. Clinical risk factors modeled included maximum oxygen requirements, peak and discharge d dimer levels, CRP levels at the onset, peak and discharge, receipt of disease-specific treatment (monoclonal antibody, dexamethasone, antibiotics), and discharge disposition.

## RESULTS

A total of 52 charts were reviewed based on inclusion criteria set in Slicer Dicer in the Epic Electronic Medical Record system. Of these, only 28 charts met the inclusion criteria. 14 patients received remdesivir, while 14 did not receive remdesivir. Baseline characteristics were similar between the two groups (Table 1). Most of the patients were Caucasian and females. The mean age of the entire group was 65.28 ± 14.14 years. The most common comorbidities were diabetes mellitus and hypertension. 39% of the patients had a history of arrhythmia, and 50% were on anticoagulation.

**Table 1:** Baseline characteristics of study population.

|  | **NON**<br>**REMDESIVIR**<br>**(n=14)** | **REMDESIVIR**<br>**(n=14)** | **TOTAL**<br>**(n=28)** |
| --- | --- | --- | --- |
| **Mean Age – yr.** | 66.57 (± 14.94) | 64.00 (± 13.80) | 65.28 (± 14.17) |
| **Age Groups in years - n, (%)** |  |  |  |
| • 18-40 | 1 (7) | 1 (7) | 2 (7) |
| • 41-64 | 4 (28) | 3 (21) | 7 (25) |
| • Above 65 | 9 (64) | 10 (71) | 19 (68) |
| **Females – no. (%)** | 7 (50) | 8 (57) | 15 (53) |
| **Race or ethnic group – no. (%)** |  |  |  |
| White or Caucasian | 6 (43) | 7 (50) | 13 (46) |
| Hispanic | 4 (28) | 4 (28) | 8 (28) |
| Black or African American | 2 (14) | 1 (7) | 3 (11) |
| Other | 2 (14) | 2 (14) | 4 (14) |
| **Tobacco use – no. (%)** | 6 (43) | 7 (54) | 12 (43) |
| **Diabetes mellitus – no. (%)** | 10 (71) | 9 (64) | 18 (64) |
| **Hypertension – no. (%)** | 14 (100) | 13 (93) | 27 (96) |
| **Coronary artery disease/peripheral**<br>**vascular disease – no. (%)** | 5 (36) | 7 (50) | 12 (43) |
| **Congestive heart failure – no. (%)** | 5 (36) | 7 (50) | 12 (43) |
| **History of lung disease– no. (%)** | 6 (43) | 5 (36) | 11 (39) |
| **Obesity (BMI>30 kg/m<sup>2</sup>) – no. (%)** | 4 (28) | 5 (36) | 9 (32) |
| **Immunosuppressed – no. (%)** | 1 (7) | 1 (7) | 2 (7) |
| **Arrhythmia – no. (%)** | 5 (36) | 6 (43) | 11 (39) |
| **Treatment – no. (%)** |  |  |  |
| Dexamethasone | 6 (43) | 14 (100) | 20 (71) |
| Antibiotics | 7 (50) | 9 (64) | 16 (57) |
| Monoclonal antibody | 1 (7) | 4 (28) | 5 (18) |
| **Anticoagulation** | 6 (43) | 8 (57) | 14 (50) |

Table 2 demonstrates no statistical difference in initial, peak, and discharge CRP levels between the two groups. D dimer levels tended to be higher in these patients, although there was no significant difference in peak and discharge d dimer levels between the two groups. The mean length of stay in the remdesivir group was 14.1 ± 9.23 days compared to 10.4± 6.4 days in the non-remdesivir group.

**Table 2.** Differences in CRP, D-dimer, Length of Stay and Disposition between Remdesivir and Non-Remdesivir treatment groups.

|  | **NON**<br>**REMDESIVIR**<br>**(N=14)** | **REMDESIVIR**<br>**(N=14)** | **P-value** |
| --- | --- | --- | --- |
| **CRP (Mean ± SD)** |  |  |  |
| Initial | 135 ± 83.8 | 122 ± 85 | 0.699 |
| Peak | 176 ± 80.3 | 174 ± 80.5 | 0.944 |
| At discharge | 77.2 ± 47 | 56 ± 64 | 0.432 |
| **D dimer (Mean ± SD)** |  |  |  |
| Peak | 896 ± 585 | 1453 ± 1529 | 0.227 |
| At discharge | 571 ± 222 | 898 ± 1023 | 0.264 |
| **Max O2 requirement**<br>**(L/min)** | 18 ± 20.6 | 15.5 ± 19.9 | 0.751 |
| **Disposition – n (%)** |  |  |  |
| Home | 3 (38) | 5 (62) | 0.723 |
| Skilled Nursing Facility | 5 (50) | 5 (50) | - |
| Died | 6 (60) | 4 (40) | 0.751 |
| **LOS in days (Mean ± SD)** | 10.4 ± 6.4 | 14.1 ± 9.23 | 0.228 |
| **Use of BiPAP – n, %** | 2 (33) | 4 (67) | 0.357 |
| **Mechanical ventilation – n, %** | 3 (60) | 2 (40) | 0.621 |
| **Hypotension – n, %** | 5 (38) | 8 (62) | 0.255 |

Maximum oxygen requirement in the remdesivir group was 15.5 ± 19.9 L/min compared to 18 ± 20.6 L/min in the non-remdesivir group. 28.5% of the patients were discharged home, although 35% of the patients died. The use of bi-level positive airway pressure (BiPAP) and mechanical ventilation was similar in the two groups. 62% of the patients in the remdesivir group were hypotensive during their hospital stay compared to 38% in the non-remdesivir group, although this was not statistically significant.

## DISCUSSION

Our study demonstrated no clinically significant difference in length of stay, oxygen requirements, or mortality in COVID-19 patients with ESRD in the remdesivir group compared to the non-remdesivir group. 64.3% of the patients were discharged from the hospital, and the mean length of stay of the group was 12.21 days. No patients’ treatment was interrupted due to hepatotoxicity. None were on peritoneal or home dialysis. Previously, only case series have been reported on the safety of remdesivir in COVID-19 patients with ESRD. To our knowledge, this is the first retrospective study to evaluate the benefit and outcomes of remdesivir in this patient group.

Remdesivir is a broad-spectrum anti-viral drug and an inhibitor of the viral RNA-dependent RNA polymerase. Once it enters the cell, it is rapidly converted by esterases into different nucleoside metabolites. Intracellular remdesivir prodrug is rapidly converted into its metabolite GS-704277 and subsequently into the monophosphate that is finally converted into the active triphosphate form. Dephosphorylation of the monophosphate form produces the remdesivir nucleoside core (GS-441524), which becomes the main circulating metabolite. The nucleoside metabolites compete with adenosine triphosphate for incorporation into viral RNA, causing premature chain termination and inhibition of viral replication. GS-441524 has potent in vitro inhibitory activity against the Middle East respiratory syndrome (MERS) coronavirus, SARS-CoV1, and SARS-CoV2. The plasma half-life of remdesivir is short (1–2 hours), but the half-life of the active metabolite GS-441524 is approximately 20–25 hours [6,7].

The other component of remdesivir includes SBECD, which increases the solubility of remdesivir. Each 100 mg of lyophilized powder and solution of remdesivir contain 3 and 6 g of SBECD (maximum recommended threshold dose 250 mg/kg per day) [8]. Animal studies have shown that accumulation of SBECD may cause renal and hepatic toxicity at doses 50 to 100 times higher than the current patients’ exposure during a 5-to-10-day cycle of remdesivir [9, 10]. SBECD remains in an ionized state after glomerular filtration and does not undergo significant tubular reabsorption. Renal excretion of a dose of remdesivir is approximately 10% as unchanged drug and 49% as GS-441524. In a case series by Davis et al., remdesivir’s half-life in ESRD patients was noted to be twice as long as in healthy volunteers. Besides, a decline in remdesivir concentration was accompanied by a simultaneous increase in GS-441524 levels, indicating accumulation. Despite this, GS-441524 appeared to be removed by HD, with post-HD concentrations 45%–49% lower than pre-HD levels [11].

Our results are similar to findings from previous studies. A case series was reported on 46 patients with acute kidney injury or chronic kidney disease that received remdesivir. Of these 46 patients, 16 (17.4%) patients had ESRD, although 36 (78.2%) patients were on hemodialysis. The authors reported that 24 (52.2%) patients were discharged from the hospital, and remdesivir caused no significant renal function abnormalities [12]. In our study, 18 (64.3%) patients were discharged from the hospital, implying similar survival rates. Estiverne et al. reported a case series on 18 patients with CrCl<30 ml/min/1.73m^2^ or on renal replacement therapy that received remdesivir. Mean CRP and d dimer levels were 133.1 mg/dl and 2196 ng/ml, comparative to our results [13]. 35% of the patients in our study died, slightly higher than prior studies that showed mortality rates of 26-32% [14, 15].

Our study has several limitations. The sample size was small and may not have been powered adequately to detect a difference. All of our patients were on intermittent HD. It is difficult to ascertain the benefit and safety of remdesivir in patients with a history of kidney transplant or chronic kidney disease stage 4 or 5 that are not on dialysis. Even though there was a baseline difference in the remdesivir group that received dexamethasone, there was no significant change in CRP levels, length of stay, mortality, or d dimer levels. However, it is unclear if the higher concurrent use of dexamethasone in the remdesivir group eclipsed potential differences in any of the outcomes. Some patients in the non-remdesivir group did not receive dexamethasone as they presented earlier in the year when the use of dexamethasone was not approved. For the same reason, some of these patients also received convalescent plasma, hydroxychloroquine, and azithromycin.

In conclusion, the addition of remdesivir to dexamethasone to treat COVID-19 in patients with ESRD had no significant effect on length of stay, oxygen requirements, mortality, CRP, and d dimer levels. Further studies are needed to study the effects of remdesivir in patients with CKD stage 4 or 5 that are not on HD. Key issues to be elucidated include the effect on renal function and the risk-benefit of a 3-day, 5-day, and 10-day regimen. Large-scale studies are also needed further to determine remdesivir’s role in this vulnerable, high-risk population.

## Data Availability

All the data is available within the manuscript.

## DECLARATIONS

### Availability of data and material (data transparency)

All the data and material is available within the manuscript.

### Code availability (software application or custom code)

Not applicable.

## Authors’ contributions

All authors contributed to the study conception and design. Conceptualization: [Kwame Dapaah-Afriyie]; Methodology: [Atin Jindal]; Formal analysis and investigation: [Muhammad Baig]; Writing-original draft preparation: [Vijairam Selvaraj]; Writing -review and editing: [Arkadiy Finn]; Supervision: [George Bayliss]. All authors read and approved the final manuscript.

### Ethics approval

The Institutional Review Board of the hospital approved the study.

### Consent to participate

Not applicable.

### Consent for publication

Not applicable.

